# Needs of key stakeholders to make advance care plans and advance directives for people with dementia – A scoping review

**DOI:** 10.1101/2025.06.30.25330415

**Authors:** Rasita Vinay, Andrea Ferrario, Sophie Gloeckler, Nikola Biller-Andorno

## Abstract

**Background:** Advance care planning (ACP) and advance directives (AD) are tools for supporting person-centered decision-making. In dementia care, the progression of cognitive decline, complex family dynamics and variability in healthcare systems pose unique challenges to effective ACP/AD implementation for people with dementia (PWD).

**Methods:** We conducted a scoping review of the literature related to ACP/AD in dementia care between 2014-2024. Studies were screened and thematically analyzed to identify current approaches, gaps and recommendations for dementia-specific ACP/AD. We identified key stakeholders involved in decision-making and highlighted procedural components for ACP/AD according to stakeholder groups.

**Results:** Forty studies were included. Key stakeholders included healthcare professionals (HCPs); family members and caregivers; PWD; dyads (PWD and their caregivers); the broader public; policymakers; and researchers. Prominent findings included: the role and training of HCPs; educational and decision-support needs; early and ongoing engagement of PWD; development and evaluation of dementia-specific tools; ethical and procedural challenges in end-of-life decision-making; and the importance of outreach and cultural sensitivity. Promising interventions include structured communication models, psychoeducational programs and tools, although few have been fully adapted for dementia.

**Conclusion:** Dementia-specific ACP/AD require a relational, flexible and ethically grounded approach that evolves with the individual’s condition. While ACP/AD should reflect the autonomous preferences of the PWD, during late-stage dementia, shared decision-making becomes central to providing care that aligns with the person’s goals and preferences. Future research should focus on inclusive tools and training; timing and process facilitation; and public health strategies to improve access and equity.

## INTRODUCTION

Dementia is a progressive condition affecting various cognitive functions, such as memory, language, thinking and decision-making [1]. As dementia advances, individuals often lose the capacity to express their care preferences, including preferences regarding living arrangements, medical treatment and end-of-life decisions [2,3]. Given this, early engagement in dementia care planning is critical to ensure that future care aligns with an individual’s values and wishes. Advance care planning (ACP) and advance directives (AD) allow individuals to consider and document their healthcare preferences and appoint a surrogate decision-maker before they lose decision-making capacity [4,5]. These interventions are particularly relevant in the case of dementia, where the person can anticipate steadily declining decision-making capacity as a result of the disease progression.

ACP/AD can help reduce uncertainty, improve outcomes, and provide guidance for families and healthcare professionals when critical care decisions arise [6]. For surrogate decision-makers such as family members and formal caregivers, ACP/AD capture the officially expressed wishes of the person with dementia (PWD) to guide decision-making on their behalf [7]. Healthcare professionals (HCPs), including physicians and nurses, contribute to interpreting these documents and applying them to treatment decision-making by providing medical guidance and safeguarding ethical and legal standards [8]. Policymakers influence the broader systems in which ACP/AD are implemented, by setting legal frameworks, directing healthcare funding, and determining institutional protocols [9].

Planning future care is first and foremost the right of the PWD. Involving multiple stakeholders can improve the efficacy of care planning in dementia and is often, eventually, necessary, but it also introduces certain challenges. Different groups bring divergent priorities, expectations, and emotional responses to the decision-making process. For example, family members may prioritize comfort and familiarity [10], whereas HCPs often emphasize clinical outcomes, safety, and resource limitations [11,12]. These views may, at times, diverge from the PWD’s previously expressed values and preferences, and progressing dementia often limits the person’s ability to self-advocate. There is a risk that the opinions and interests of others may supersede those of the PWD, especially when the PWD’s wishes are not clearly documented or are interpreted differently by different stakeholders. PWD often also experience coercion. ACP/AD can serve as important safeguards.

Despite the recognized importance of ACP and AD in dementia care, there remains no clear consensus on which procedural components, e.g., interactions, tools, and policies, best meet the unique needs of PWD. A central challenge lies in tailoring these directives and processes to the progressive and variable nature of dementia. PWD have distinct cognitive needs that influence their ability to participate in ACP/AD over time. These evolving impairments demand ACP/AD approaches that are both flexible enough to accommodate changing capacities and specific enough to guide care in later stages. As such, dementia-specific tools are essential to ensure that planning processes remain meaningful, inclusive and responsive to the unique trajectory of dementia. This scoping review addresses the question *“What are the critical areas of consideration for individuals making advance care decisions in dementia care?”* by identifying (1) key stakeholders, and (2) the procedural components required to support their needs and roles while engaging with ACP/AD completion. As a result, this would allow us to determine how best to design tools that are tailored to PWD. By clarifying what makes ACP/AD more effective and efficient and to whom within dementia care, we aim to contribute to more structured and ethical approaches to dementia care planning.

## METHODS AND MATERIALS

### Overview

The present scoping review was conducted following the methodological framework proposed by Arksey and O’Malley [13] and reported according to the PRISMA-ScR (Preferred Reporting Items for Systematic Reviews and Meta-Analyses Extension for Scoping Reviews) checklist **(Supplemental material 1)**. The protocol for the review was preregistered on the Open Science Framework registry in January 2025 [14].

### Search strategy

A comprehensive list of specific terms relating to care planning and dementia were identified using a web-based search of relevant published research. A search string was developed for databases: Pubmed, EMBASE, Scopus and Cochrane, and a comprehensive search performed first on 09.04.2024 and updated on 31.10.2024. The detailed search strategy and search strings can be found in **Supplemental material 2**.

### Eligibility Criteria

Articles were screened following specified inclusion and exclusion criteria established by all authors. For inclusion, articles must significantly discuss components of ACP or AD within dementia care and must be original research articles. Articles from 2014-2024 were included to capture timely and relevant findings. Articles were excluded if they did not have a specific focus on dementia, ACP, or AD. Editorials, commentaries, book front/back ends, conference proceedings/abstracts, literature reviews, preprints, protocols and articles not in English were also excluded.

The choice to exclude review articles was deliberate, due to the risk of duplicating results and over emphasizing certain findings as the reviewed articles would already be included individually.

### Screening, Data Extraction and Analysis

Screening for articles was conducted in two stages: (1) title and abstract screened by one reviewer (RV), (2) full-text review conducted by three reviewers independently (RV, AF and SG). An interrater reliability of 94.38% was obtained, where any disagreements were solved through discussion.

After full-text review, reviewers RV and AF performed data extraction of all eligible articles. Data extracted included: title of article, DOI, authors, year of publication, country of study, aims, study type, study population, inclusion/exclusion criteria of the study itself, sample size, study setting, intervention (if applicable), outcomes measured, key results, focus on AD or ACP and key considerations for decision-making.

A thematic analysis based on full-text review of the included studies was conducted by RV and further consolidated by AF. The thematic analysis extracted content related to key considerations in advance care planning for dementia care.

## RESULTS

### Search findings

The database search retrieved 6,921 records, of which 2,640 were removed as duplicates. 4,281 records were screened by their title and abstract, of which 180 were determined eligible for full-text review. Of these, 138 articles were excluded and 2 could not be retrieved, resulting in a final pool of 40 reports (see **Figure 1)**.

**Figure 1.**
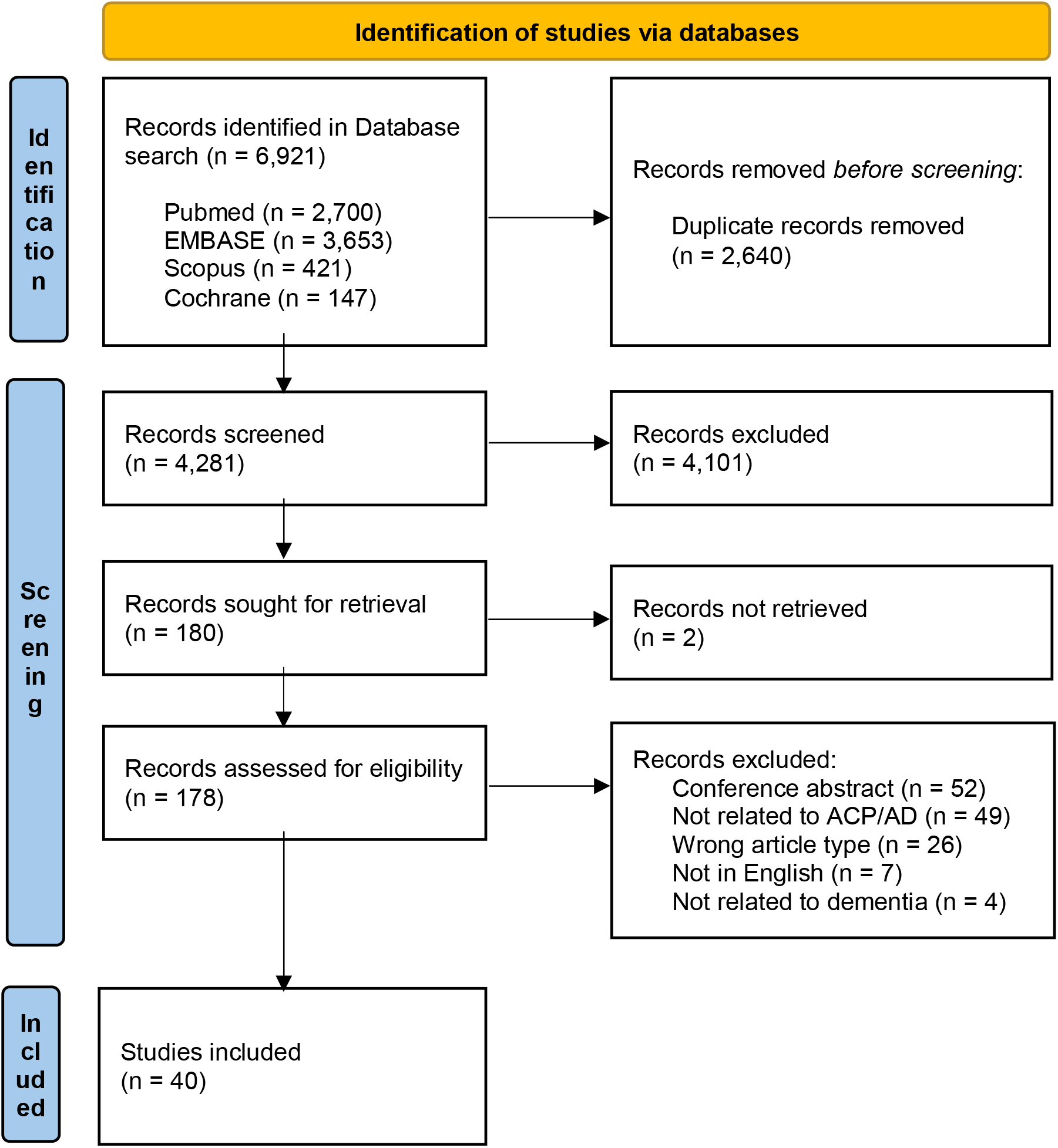
PRISMA flowchart showing the study selection. *Notes: ACP: advance care plan; AD: advance directive*.

### Characteristics of included studies

In this scoping review, 16 of the included studies were conducted in the USA, five in Belgium, three in Germany and Taiwan, two in Australia, Canada and Finland, and one study each in Hong Kong, Israel, Netherlands, New Zealand, Singapore, Switzerland and the United Kingdom. 19 studies were published from 2014-2019 and 21 studies from 2020-2024.

Of the included studies, 13 were qualitative studies, six were cross-sectional studies, four were cluster RCTs, four were mixed methods studies, four were pilot studies, three were pre-test/post-test interventions, two were retrospective studies, and there was one consensus development, experimental, quasi-experimental and user-centered design study each.

Twenty-five studies focused primarily on ACP, eight on both ACP and AD and seven strictly on AD. A summary of the included studies can be seen in **Supplemental material 3**.

### Key considerations in dementia care

Based on the included studies, we conducted a thematic analysis structured as follows. First, we identified relevant stakeholders involved in decision-making in dementia care: healthcare professionals (HCPs), caregivers and family members, PWD, dyads (PWD and their caregivers), as well as public, policy and research stakeholders. This specific focus on prioritizing stakeholders allows us to identify how ACP/AD completion takes place across different care contexts by highlighting the distinct roles, needs and interactions of PWD and those supporting their decision-making. Next, we extracted themes specific to each stakeholder group through inductive coding. Data segments were coded iteratively through emerging themes, where three rounds of coding were completed. Two reviewers (RV and AF) went through the final codes and themes of the analyzed studies to ensure reliability and consistency. **Table 1** presents these themes along with illustrative examples from the reviewed studies.

**Table 1.**
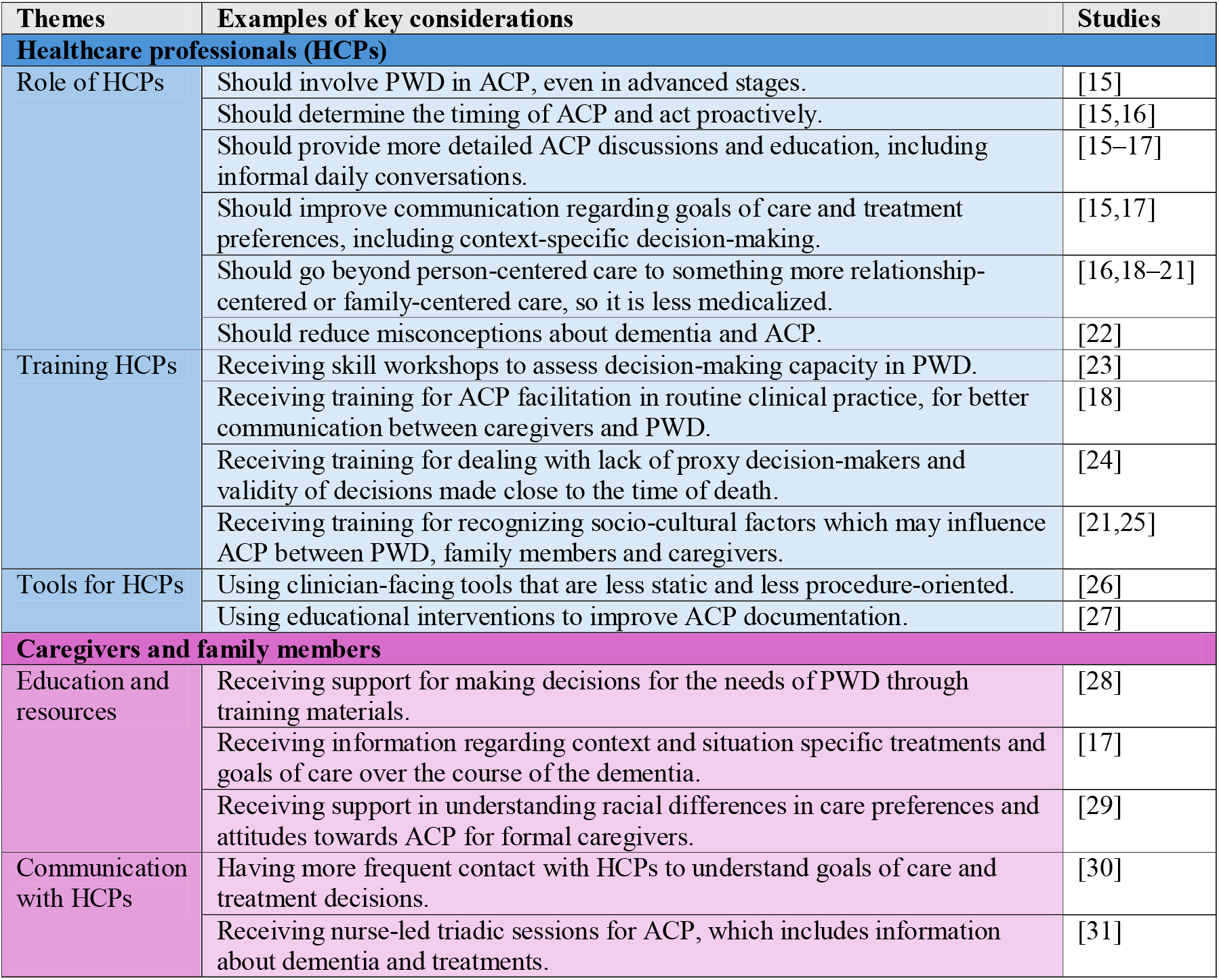
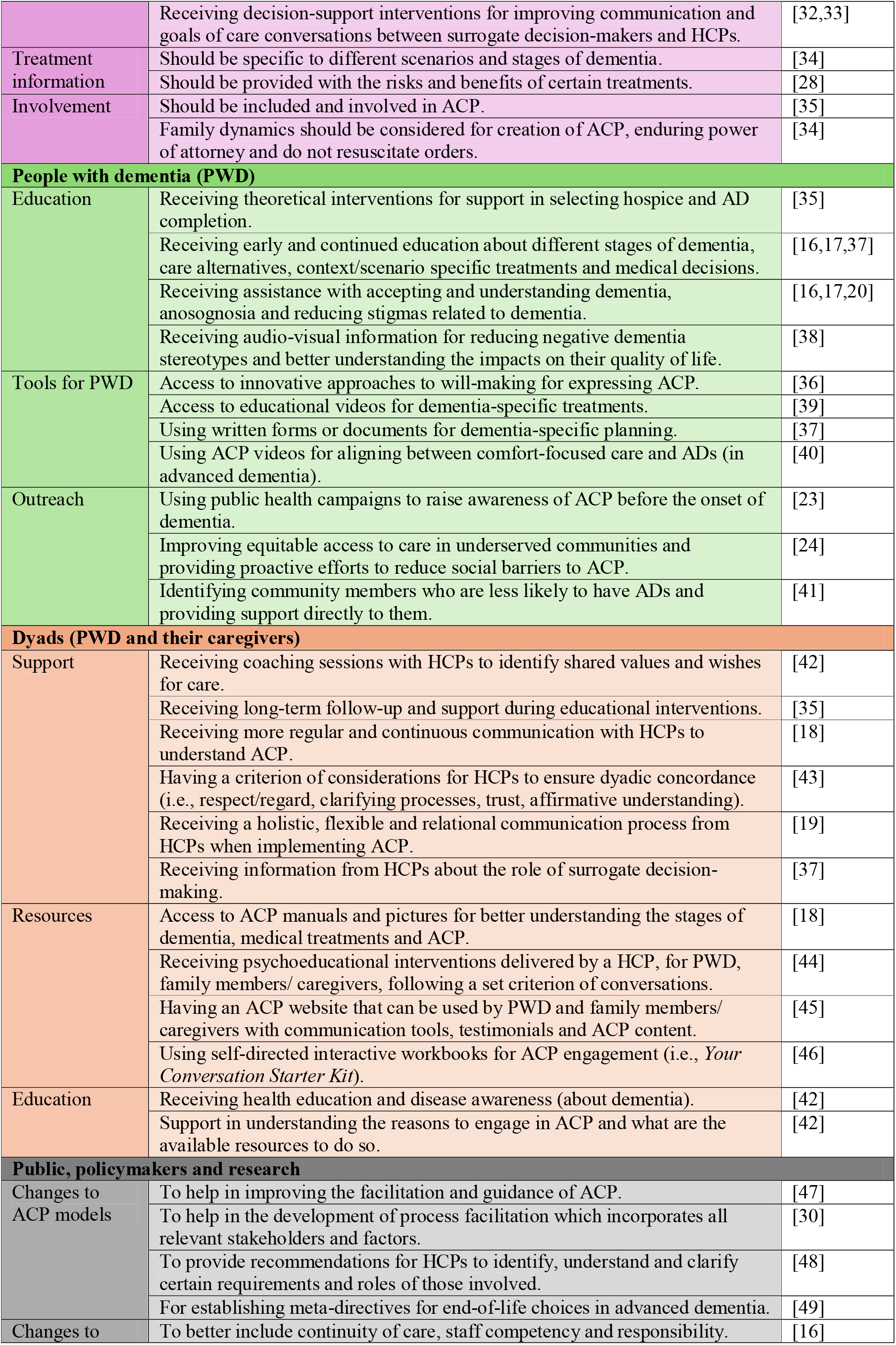
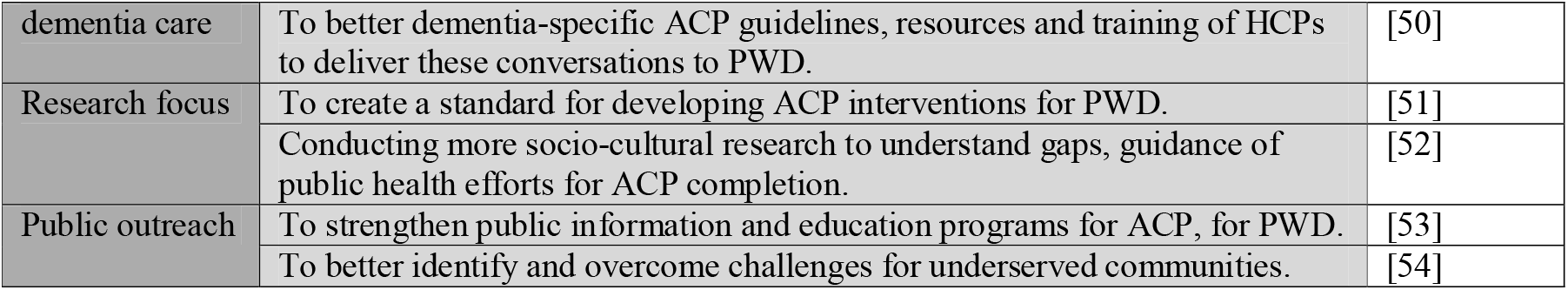
Key considerations of the relevant stakeholders in dementia care.

## DISCUSSION

### Principal findings

This scoping review identified dementia-specific procedural components of ACP and AD that are relevant factors for decision-making in such cases. Across the 40 included studies, ACP and AD were explored in relation to HCPs; caregivers and family members; PWD; dyads (PWD and their caregivers); the public; policymakers; and researchers. Key findings highlight the value of structured and consistent ACP, including the use of AD; the specific role of HCPs in facilitating decision-making and how to better prepare them for this role; and the need for resources that support both caregivers and PWD in making informed care and treatment decisions. The following sections provide a more in-depth analysis of findings and implications for future ACP/AD completion.

### Key considerations for healthcare professionals (HCPs)

#### Role of HCPs

HCPs play a vital role in ensuring that PWD are included in ACP/AD decision-making, even in advanced stages of dementia [15]. Their involvement helps reduce misconceptions about dementia or ACP/AD [22] and supports ethical, person-centered care. Meaningful engagement in ACP/AD depends on timely discussions, ideally during admission (particularly in the first weeks/months), during crises and through ongoing informal daily conversations [15,16]. During these discussions, it is important for HCPs to provide more information, especially towards helping to clarify specific goals of care and treatment preferences, such that [17]. Finally, a shift towards relationship-or family-centered care may also be necessary, as overly medicalized individualistic approaches may limit participation and shared understanding [16,18–21].

#### Training of HCPs

Many HCPs struggle with assessing a patient’s decisional capacity and navigating ethically complex scenarios. Targeted training, such as skill-development workshops or practice through role play, as well as having tools to use with patients and loved-ones can improve confidence and communication [23] Skills for facilitating discussions, incorporating ACP into the routine clinic pathway and engaging in regular communication with PWD and their caregivers are especially important and could help reduce decisional conflicts in ACP/AD planning [18]. Practice addressing ethically complex scenarios, such as the absence of proxy decision-makers or g navigating decisions made close to the time of death where documented ADs may not reflect the patient’s self-formulated wishes due to the timing of when they were documented,[24] could also be improved through targeted training provided to HCPs. Furthermore, socio-cultural and contextual factors may influence ACP/AD completion between PWD, family members and caregivers [21,25]; thus, it may be important for HCPs to receive cultural competency training, related to dementia.

#### Tools for HCPs

Current ACPs are often unable to capture the evolving needs of PWD. Therefore, clinician-facing tools should be enhanced specifically for dementia to better enable ACP for the needs of PWD, where they explore values and explore the caregiver role, which can be done by making the ACP procedure less static and procedure-oriented [26] Educational interventions or decision tools for HCPs could help with improving ACP documentation and communication [27], however the impact of such tools depends on the effectiveness of integrating them into routine care.

In summary, future research should explore how HCPs can be effectively supported in initiating and sustaining ACP conversations with PWD, particularly in the early stages of care and during key transition points such as admission or crisis events. There is a need to develop and evaluate dementia-specific training programs and tools that equip HCPs to navigate the complexities of decision-making capacity, cultural sensitivities, and family dynamics in ACP and AD planning. Further studies should investigate the design and implementation of procedure-oriented ACP tools that move beyond procedural checklists and instead emphasize the evolving values, preferences and social context of PWD and their caregivers [55].

### Caregivers and family members

#### Education and resources

Caregivers and family members are often central to making decisions for PWD, yet many lack the knowledge and support to confidently participate in ACP/AD planning [28]. Here, tailored educational resources are essential [47], ideally including scenario-based guidance that captures the progression of dementia and evolving goals of care [17]. These materials should help caregivers navigate complex treatment decisions over time. For formal caregivers, training should also address cultural variations in care preference, including racial and ethnic attitudes towards ACP/AD and end-of-life decisions [29].

#### Communication with HCPs

Regular and meaningful communication with healthcare professionals helps caregivers and family members to better understand medical decisions and the broader goals of care [30]. Studies highlight the potential of decision-support interventions to improve this dialogue, especially in facilitating difficult conversations between surrogate decision-makers and clinical teams [32,33]. Caregivers and family members also reported feeling more engaged when ACP/AD sessions are led by nurses and take place in triadic settings, where the PWD, carers and HCP are all present [31]. These formats foster trust and shared understanding and may reduce the sense of isolation caregivers often report in decision-making processes.

#### Dementia treatment information and involvement in care planning

Providing information that is tailored to the stage and context of dementia is critical [34]. Caregivers need clarity around the potential risks and outcomes of specific treatments, as well as the practical and ethical implications of decisions such as resuscitation orders or feeding interventions [28,36]. Involving caregivers and family members in the planning process acknowledges the importance of relational dynamics and promotes decisions that are aligned with the values of both the PWD and their support network [35]. Identifying an enduring power of attorney or health proxy should also be approached as a collaborative and anticipatory process, not just a legal formality [36].

In summary, future research should examine how triadic decision-making models - engaging HCPs, patients and families - used in other chronic illnesses, can inform dementia care planning. Such models could help overcome the challenge of medicalization in dementia-related ACP by fostering trust and shared understanding among all parties involved.

### People with dementia (PWD)

#### Education

Educational support is essential to help PWD better understand their diagnosis, care options and the different medical decisions that they may face [16,17,20,37]. Early and on-going education can improve acceptance of the condition, reduce anosognosia and prepare individuals for future care planning [17]. Interventions using audio-visual formats, such as videos about living with dementia, may help reduce stigma, support quality of life discussions and introduce relevant resources [38]. There may be a need for more theory-based educational interventions that guide PWD in considering hospice care and how to complete ADs [35].

#### Tools for PWD

PWD have expressed interest in more accessible and innovative approaches, such as through digital tools, to write wills that include their ACP/AD [36]. Other innovative approaches may include multimedia and plain-language tools (i.e., like the *PREPARE* tool [55]), that help users clarify their values, designate proxies and more clearly express care preferences. These tools also help family members in understanding and following the wishes of the PWD. Dementia-specific resources, such as educational videos [39], or structured forms and documents tailored to treatment contexts [37], can make ACP more actionable. For those in advanced stages of dementia, visual aids showing comfort-focused care options may help align decisions with documented ADs [40].

#### Outreach

A few studies highlight the necessity of supporting outreach around advance care of PWD. For instance, public health campaigns can raise awareness and normalize early planning, particularly before cognitive decline begins [23]. Community-based strategies are needed to reduce social and structural barriers to care planning, especially in underserved communities where ACP/AD engagement is low [24]. Proactive efforts to identify and support community members who are less likely to complete care plans, including culturally targeted outreach and local advocacy, could promote more equitable access [41].

In summary. future research should explore early and continuous educational interventions that help PWD understand their diagnosis, care options and medical decisions. There is a need to develop dementia-specific, digital and context-sensitive tools, such as videos or interactive documents that support PWD in expressing advance care preferences, ideally in collaboration with caregivers. Studies should also assess the effectiveness of outreach strategies, including public health campaigns and community-based efforts, in increasing awareness and equitable access to ACP. Finally, targeted interventions are needed to reach underserved populations who are less likely to engage in ACP/AD.

### Dyads (PWD and their caregivers)

#### Support

Although shared decision-making can support collaborative care planning, it is important to distinguish it from advance decision-making, which prioritizes the autonomy of the person while they still have capacity. However, as the capacity of a PWD declines, shared decision-making is best supported through structured coaching with a HCP where sessions help clarify values and future care preferences [42]. Such interventions should include regular follow-ups to support ongoing communication [18]. Dyads often benefit from guidance on the surrogate’s role and the extent of decision-making leeway they may have [37]. Educational programs aimed at dyads should include long-term support, with content tailored to relational dynamics, achieving dyadic concordance and the evolving nature of dementia [43]. This is particularly important for dyads affected by young-onset dementia, where care planning should especially be holistic, flexible and relational rather than procedural [19].

#### Resources & Education

A range of tools and resources have been developed to support dyads in engaging with ACP/AD, including visual aids [18], interactive websites [45], physical resources [46] and psychoeducational interventions [44]. ACP manuals and pictures can clarify life-sustaining medical procedures and help dyads prepare for end-stage dementia [18]. Online platforms offer structured content, peer testimonials and interactive features, but should allow users to navigate at their own pace [45]. Self-directed workbooks, like *Your Conversation Starter Kit*, support early ACP engagement by prompting reflection on quality of life, family involvement in decision-making and timing of conversations [46]. Further, psychoeducational interventions delivered by an interventionist, such as *SPIRIT* [44], which combine education, goal setting and planning support, have also been effective in helping dyads align on goals of care [41].

Overall, dyads require relational, flexible and sustained support to meaningfully engage in dementia-related ACP. Future research should examine how existing tools like *PREPARE* [55] could be adapted and tested for dyads. Specifically, health education and disease awareness interventions should be tailored to promote joint reflection and shared understanding of the need to engage in ACP processes together, i.e., early in the dementia, providing resources for within the community and contingency planning [42].

### Public, policy and research

#### Changes to ACP models and dementia care

Literature suggests several changes to improve current ACP models. These include better facilitation by HCPs [47] and clearer guidance to ensure all aspects of the ACP process are addressed [30,48]. When it comes to end-of-life care, or decisions during late-stage dementia, it would be important to include all relevant stakeholders in shared decision-making, to ensure decisions are made according to the PWD’s previous autonomous wishes and current medical assessments. For instance, there is a known challenge when ADs prohibiting life-supporting treatment in late-stage dementia are not followed if the PWD later appears to be “happy” in their dementia [49]. In such conflicts, implementing an explicit meta-directive in the AD, which provides documented instructions for addressing these cases may offer a solution [49].

A synthesis of key recommendations for ACP models includes the following considerations [30,48]:

1. *Physician’s strategy and ACP initiation:* Define timing, whom to involve, discussion topics and context-specific elements.
2. *Patient assessment:* Evaluate the patient’s condition and mental capacity.
3. *ACP conversations with PWD:* Use communication strategies that involve surrogates, increase disease awareness and explore experiences, needs, fears and care preferences.
4. *Clarify roles of supporters:* Understand the willingness, degree of involvement and needs of family members/ caregivers, while promoting consensus within the care network.
5. *Non-verbal ACP inclusion:* Maximize participation of PWD by attending to non-verbal cues, behaviors and emotions, with caregiver involvement.
6. *Documentation of preferences:* Record values, care goals, AD and legal proxies; ensure ACPs are regularly reviewed and communicated to care teams.
7. *End-of-life decision-making:* Balance previously expressed wishes with current best interests, in consultation with the broader care team.
8. *Policy support:* Develop national frameworks that include HCP training, implementation support and organizational integration of ACP.

Beyond ACP models, restructuring dementia care models more broadly is also needed. This includes clearer care pathways to ensure a continuity of care, role definitions, guidance for ACP implementation, support for professional responsibilities and addressing staff shortages and resource limitations [16,50].

#### Research focus

This review identified a limited focus on intervention development [48], with no existing standards for ACP-based dementia interventions [51]. More targeted research is needed to develop, test and refine these tools. In addition, socio-cultural research should explore how ACP is influenced by social norms, beliefs and health system structures [52]. Comparative studies within a country, as well as between countries, can help tailor interventions to different populations, [49], reduce stigma and support more inclusive public health strategies [52].

#### Public outreach

Public education remains a critical factor in increasing engagement with ACP [53]. Strengthening outreach in underserved communities requires more than information alone, but rather relationship-building and community participation. Effective strategies include outreach health programs such as affordable medical clinics in underserved neighborhoods, partnering with trusted local organizations and forming community advisory boards to support culturally relevant outreach [54].

### Unaddressed areas within this review and future research avenues

This review highlights that while there is growing recognition of the importance of ACP/AD in dementia care, several gaps remain that limit the effective implementation. Limited public and professional understanding of ACP/AD, unclear roles for surrogate decision-makers and delays in initiating these conversations all contribute to fragmented planning. Furthermore, decision-making is often led by caregivers, especially under high levels of burden, which may inadvertently clash with the wishes of PWD. It should be clarified that while this review highlights the need to initiate dementia-specific ACP/AD completion related activities as soon as dementia is diagnosed, it does not provide evidence in how to initiate conversations, frequency of review or care topics.

Certain questions also could not be answered by this review due to a lack of evidence, including the need for capacity reviews and assessments for cognitive decline – both of which should form necessary considerations for ACP/AD completion. Additionally, the lack of dementia specific ACP/AD forms/tools was striking. While our review emphasizes current effort in educational interventions, future research avenues need to specify what makes dementia-specific ADs a good vehicle for capturing the evolving preferences, values and treatment boundaries of individuals across different stages of cognitive decline. Future research should focus on identifying structural, ethical and contextual components that make dementia-specific ADs clear, actionable and sensitive to the progressive nature of dementia. Co-design approaches that involve PWD, caregivers and HCPs, are especially promising to ensure tools are practical and sensitive to users’ needs.

Designing dementia-specific ACP/AD tools also requires grappling with ethical and legal uncertainties surrounding decisional capacity, influence, and temporal consistency. As in legal assessments of testamentary capacity [56], PWD may experience fluctuating cognitive function, raising critical questions about when and how capacity should be evaluated, and whether a minimal threshold should be defined for making binding decisions [57,58]. Safeguards, similar to those used to detect undue influence in financial wills, may be needed to ensure that ACP/AD reflect the individual’s authentic wishes rather than those of others [56,59]. Moreover, ethics literature highlights the tension between prior autonomous choices and current experiential interests, prompting the need for ACP/AD formats that anticipate such conflicts and clarify how the person would want these to be reconciled [60–62]. This calls for dementia-sensitive communication strategies [60] and the inclusion of care preferences that go beyond generic AD templates to address the specific trajectory and care realities of dementia.

Finally, future studies should continue to explore the timeliness and initiation of ACP conversations, building on earlier findings that such discussions often occur too late. Early, ongoing communications that are guided by accessible resources and supported by care teams, may help reduce the burden on caregivers and ensure that planning reflects the true values and preferences of PWD. To ensure ADs continue to be led by autonomous preferences of the PWD, it may also be important for future research to explore how the needs/preferences of caregivers or HCPs may influence their decision-making.

### Limitations of this scoping review

This scoping review focuses specifically on a diagnosis of dementia and did not include studies discussing mild cognitive impairments (MCI), subjective cognitive decline (SCD), or other cognitive impairments not meeting the clinical criteria for dementia. The rationale was that while MCI and SCD can be early indicators of dementia, they are distinct conditions where individuals often retain decision-making capacity, which may influence how they engage in ACP conversations. However, due to this criterion, we may have potentially missed a unique context and perspective of early interventions or techniques that may help shape care preferences before dementia progresses.

## CONCLUSION

This scoping review highlights the multidimensional nature of ACP and AD in dementia care, involving healthcare professionals, caregivers and family members, persons with dementia and broader public, policy and research frameworks. While current studies provide foundational insights into ACP/AD implementation, future research should prioritize the creation of dementia-specific, user-friendly tools and the adaptation of existing ACP-supporting frameworks for the dementia context. There is also a critical need for early, ongoing education for PWD, culturally inclusive interventions and scalable public outreach efforts to promote awareness and equitable access. Crucially, advancing ethically grounded approaches require reconciling tensions between autonomy and best-interest judgements in later stages of dementia. Future ACP models must account for this ethical complexity while remaining flexible, inclusive and responsive to relational dynamics in care settings.

## Data Availability

All data produced in the present work are contained in the manuscript.

https://osf.io/ecy4x/

## ACKNOWLEDGEMENTS

RV would like to thank the Ethox Centre, University of Oxford and the Caroline Miles Scholarship which allowed her to work on parts of this study during a research visit in November 2024.

## COMPETING INTERESTS

The authors have no competing interests to declare.

## FUNDING

This research did not receive any funding.

## Notes

### Competing Interest Statement

The authors have declared no competing interest.

### Funding Statement

This study did not receive any funding.

