## Supplemental Material 2 for "Needs of key stakeholders to make advance care plans and advance directives for people with dementia – A scoping review"

Search String [UPDATED OCTOBER 2024]

| **Database** | **Results** | **Search String** |
|---|---|---|
| Pubmed<br>09.04.2024 | 2,458 | (advance directive*[Title/Abstract] OR advance care plan*[Title/Abstract] OR end-of-life decision*[Title/Abstract] OR end-of-life plan*[Title/Abstract] OR decision-making[Title/Abstract] OR patient preference*[Title/Abstract]) AND (dement*[Title/Abstract] OR alzheimer*[Title/Abstract]) AND (2014:2024[pdat]) |
| [Updated]<br>Pubmed<br>31.10.2024 | 242 | (advance directive*[Title/Abstract] OR advance care plan*[Title/Abstract] OR end-of-life decision*[Title/Abstract] OR end-of-life plan*[Title/Abstract] OR decision-making[Title/Abstract] OR patient preference*[Title/Abstract]) AND (dement*[Title/Abstract] OR alzheimer*[Title/Abstract]) AND (2024/4/9:2024/10/31[pdat]) |
| EMBASE<br>09.04.2024 | 3,305 | ('advance directive*':ab,ti OR 'advance care plan*':ab,ti OR 'end-of-life decision*':ab,ti OR 'end-of-life plan*':ab,ti OR 'decision making':ab,ti OR 'patient preference*':ab,ti) AND (dement*:ab,ti OR alzheimer*:ab,ti) AND [2014-2024]/py |
| [Updated]<br>EMBASE<br>31.10.2024 | 348 | ('advance directive*':ab,ti OR 'advance care plan*':ab,ti OR 'end-of-life decision*':ab,ti OR 'end-of-life plan*':ab,ti OR 'decision making':ab,ti OR 'patient preference*':ab,ti) AND (dement*:ab,ti OR alzheimer*:ab,ti) AND [2024-2024]/py |
| SCOPUS<br>09.04.2024 | 379 | ( TITLE ( "advance directive*" OR "advance care plan*" OR "end-of-life decision*" OR "end-of-life plan*" OR "decision making" OR "patient preference*" ) AND ABS ( "advance directive*" OR "advance care plan*" OR "end-of-life decision*" OR "end-of-life plan*" OR "decision making" OR "patient preference*" ) AND TITLE ( dement* OR alzheimer* ) AND ABS ( dement* OR alzheimer* ) ) AND PUBYEAR > 2013 AND PUBYEAR < 2025 |
| [Updated]<br>SCOPUS<br>31.10.2024 | 42 | ( TITLE ( "advance directive*" OR "advance care plan*" OR "end-of-life decision*" OR "end-of-life plan*" OR "decision making" OR "patient preference*" ) AND ABS ( "advance directive*" OR "advance care plan*" OR "end-of-life decision*" OR "end-of-life plan*" OR "decision making" OR "patient preference*" ) AND TITLE ( dement* OR alzheimer* ) AND ABS ( dement* OR alzheimer* ) ) AND PUBYEAR > 2023 AND PUBYEAR < 2025 |
| Cochrane<br>09.04.2024 | 142 | (advance NEXT directive* OR advance NEXT care NEXT plan* OR end-of-life NEXT decision* OR end-of-life NEXT plan* OR decision-making OR patient NEXT preference*):ti OR (advance NEXT directive* OR advance NEXT care NEXT plan* OR end-of-life NEXT decision* OR end-of-life NEXT plan* OR decision-making OR patient NEXT preference*):ab AND (dement* OR alzheimer*):ti OR (dement* OR alzheimer*):ab with Cochrane Library publication date Between Jan 2014 and Mar 2024 |
| [Updated]<br>Cochrane<br>31.10.2024 | 5 | (advance NEXT directive* OR advance NEXT care NEXT plan* OR end-of-life NEXT decision* OR end-of-life NEXT plan* OR decision-making OR patient NEXT preference*):ti OR (advance NEXT directive* OR advance NEXT care NEXT plan* OR end-of-life NEXT decision* OR end-of-life NEXT plan* OR decision-making OR patient NEXT preference*):ab AND (dement* OR alzheimer*):ti OR (dement* OR alzheimer*):ab with Cochrane Library publication date Between Apr 2024 and Oct 2024 |
